# Alternative splicing in the lung influences COVID-19 severity and respiratory diseases

**DOI:** 10.1101/2022.10.18.22281202

**Authors:** Tomoko Nakanishi, Yossi Farjoun, Julian Willett, Richard J. Allen, Beatriz Guillen-Guio, Sirui Zhou, J. Brent Richards

## Abstract

Hospital admission for COVID-19 remains common despite the successful development of vaccines and treatments. Thus, there is an ongoing need to identify targets for new COVID-19 therapies. Alternative splicing is an essential mechanism for generating functional diversity in protein isoforms and influences immune response to infection. However, the causal role of alternative splicing in COVID-19 severity and its potential therapeutic relevance is not fully understood. In this study, we evaluated the causal role of alternative splicing in COVID-19 severity and susceptibility using Mendelian randomization (MR). To do so, we performed two-sample MR to assess whether *cis*-sQTLs spanning 8,172 gene splicing in 5,295 genes were associated with COVID-19 outcomes in the COVID-19 Host Genetics Initiative, including up to 158,840 COVID-19 cases and 2,782,977 population controls. We identified that alternative splicing in lungs, rather than total RNA expression of *OAS1, ATP11A, DPP9* and *NPNT*, was associated with COVID-19 severity. *MUC1* splicing was associated with COVID-19 susceptibility. Further colocalization analyses supported a shared genetic mechanism between COVID-19 severity with idiopathic pulmonary fibrosis at *ATP11A* and *DPP9* loci, and with chronic obstructive lung diseases at *NPNT*. We lastly showed that *ATP11A, DPP9, NPNT*, and *MUC1* were highly expressed in lung alveolar epithelial cells, both in COVID-19 uninfected and infected samples. Taken together, these findings clarify the importance of alternative splicing of proteins in the lung for COVID-19 and other respiratory diseases, providing isoform-based targets for drug discovery.

## Introduction

Despite the successful development of vaccines^1,2^ and treatments^3,4^, hospital admission for severe COVID-19 and long-term sequela for COVID-19 remain common^5,6^. COVID-19 is now a leading cause of death, accounting for 350,000 deaths^7^ and the estimated COVID-19 mortality rate is generally higher than that of influenza by 2 to 100-fold, which vary by the study design^7,8^. Thus, there is an ongoing need to identify mechanistic targets for therapeutic development to reduce the risk of severe COVID-19.

Using human genetics methods, several host factors have been identified to influence COVID-19 severity, including *OAS1*, type I interferon and chemokine genes^9–14^. Human genetics can provide new biological insights into disease pathogenesis and the therapeutic targets and evidence from human genetics increases the probability of drug development success^15,16^. While prior studies have evaluated causal roles of both RNA expression and circulating proteins in COVID-19 outcomes^9,12,13^, the causal role of alternative splicing has not been fully investigated.

Alternative splicing is an essential mechanism for generating functional diversity in the isoforms, through which multiple mRNA isoforms are produced from a single gene, often in tissue-specific patterns^17^. Alternative splicing has been implicated to play an important role in immune response to infections in humans^17^, and this might be the case for SARS-CoV-2 infection. The genetic determinants of alternative splicing have been identified using splicing quantitative trait loci (sQTL) studies, which indicate that splicing is under strong genetic control in humans and often has direct effects on protein isoforms. Indeed, we have recently identified that a Neanderthal-interrogated isoform of OAS1, which is strongly regulated by an sQTL, affords protection against COVID-19 severity^9^. Given this evidence, we hypothesized that alternative splicing could partially explain the variability in COVID-19 outcomes in humans.

In this study, to test our hypothesis, we undertook two-sample MR and colocalization analyses to combine results from genome-wide association studies (GWASs) of gene splicing and COVID-19 outcomes^18^. To do so, we first obtained the *cis*-regulatory genetic determinants of gene splicing (*cis*-sQTLs) in lungs and whole blood, two relevant tissues that influence acute SARS-CoV-2 infection, from the GTEx Consortium^19^. We then used MR to assess whether these gene splicing events have effects on COVID-19 outcomes using the GWASs from the COVID-19 Host Genetics Initiative^11^. Next, all findings were assessed for colocalization to ensure that the gene splicing and COVID-19 outcomes shared a common etiological genetic signal and that the MR results were not biased by linkage disequilibrium (LD). We also compared effects of alternative splicing to total RNA expression and evaluated the expression in the lung transcriptome datasets. Finally, we evaluated whether such alternative splicing is a shared pathophysiological mechanism with other respiratory diseases.

## Results

### MR using cis-sQTLs, and colocalization analyses

The study design is illustrated in **Fig. 1**. We obtained the genetic determinants of gene splicing (splicing quantitative trait locus [sQTL]) that influence alternative splicing events, which were quantified as normalized intron excision ratios, *i*.*e*. the proportion of reads supporting each alternatively excised intron, by LeafCutter^20^ (using a multiple-testing correction threshold of a false discovery rate of 5% per tissue) in GTEx v.8^19^. We chose to examine only *cis-*sQTLs, which act *cis* to the coding genes, as it is less likely that these act through horizontal pleiotropy, where the SNPs have effects on diseases (here, COVID-19 outcomes) independently of the gene splicing. We also focused on two COVID-19-relevant tissues; lungs (N=515, of which 452 were of European American ancestry) and whole blood (N=670, of which 570 were of European American ancestry) since pulmonary symptoms are the major determinant of hospital admission and the immune system plays a major role in host response to SARS-CoV-2.

**Fig. 1.**
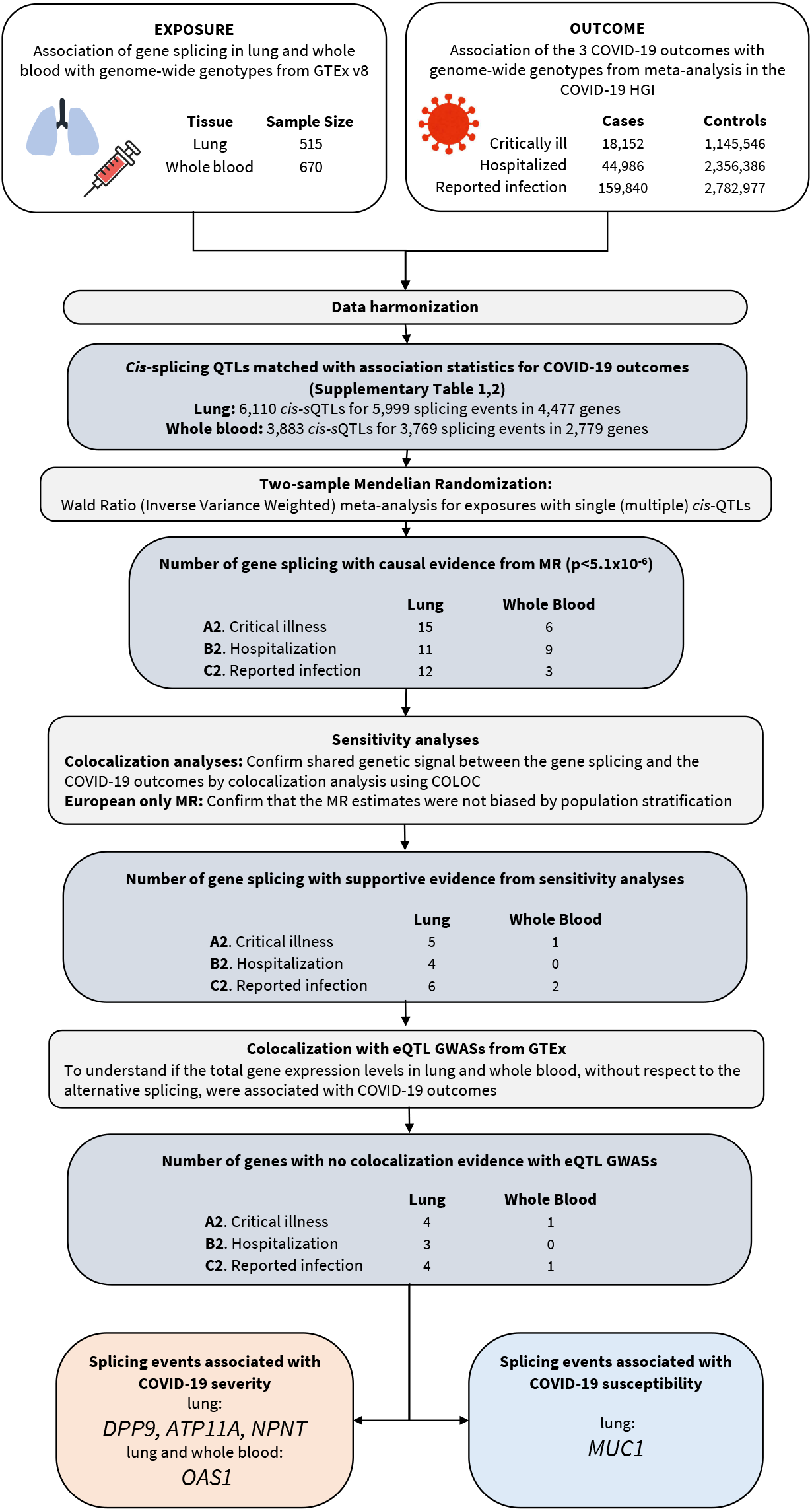
Flow Diagram of Study Design.

A total of 4,477 genes in lungs and 2,779 genes in whole blood (5,295 unique genes in total) contained conditionally independent *cis-*sQTLs that were also present the GWAS meta-analyses of COVID-19 Host Genetics Initiative^11^ release 7 (https://www.covid19hg.org/results/r7/), which included results from the GenOMICC^12^ study. We then undertook two-sample MR analyses using 6,110 *cis-*sQTLs in lungs and 3,883 *cis-*sQTLs in whole blood as genetic instruments for gene splicing for three COVID-19 outcomes: 1) critical illness (defined as individuals experiencing death, mechanical ventilation, non-invasive ventilation, high-flow oxygen, or use of extracorporeal membrane oxygenation) owing to symptoms associated with laboratory-confirmed SARS-CoV-2 infection (18,152 cases and 1,145,546 controls, of which 13,769 and 1,072,442 were of European ancestry); 2) hospitalization owing to symptoms associated with laboratory-confirmed SARS-CoV-2 infection (44,986 cases and 2,356,386 controls, of which 32,519 and 2,062,805 were of European ancestry); and 3) reported SARS-CoV-2 infection defined as laboratory-confirmed SARS-CoV-2 infection, electronic health record or clinically confirmed COVID-19, or self-reported COVID-19, with or without symptoms of severity (159,840 cases and 2,782,977 controls, of which 122,616 and 2,475,240 are of European ancestry). Our primary MR analyses used data from individuals of all ancestries, so as not to discard data for minority populations^21^, albeit the vast majority of the individuals were of European ancestry; 86.2% in GTEx and 88.8% in COVID-19 HGI). We also performed MR analyses using data derived only from European population as described below.

MR analyses identified 29 genes, whose alternative splicing in lung influenced COVID-19 outcomes. Using whole blood-derived alternative splicing we identified 20 genes influencing the same outcomes. All associations survived a p-value threshold of 5.1×10^−6^, which was based on Bonferroni correction using the number of splicing events tested (in total 8,172 [5,999 in lung and 3,769 in whole blood], **Methods, Supplementary Table 1,2**). We first replicated the association of COVID-19 outcomes with *OAS1* gene splicing which increases the excision of the intron junction at chr12:112,917,700-112,919,389 [GRCh38] both in lung and whole blood, that corresponds to an increased level of the p46 isoform, a prenylated form of OAS1 with higher anti-viral activity than the p42 isoform^22–24^ (**Fig. 2, Extended Data Fig. 1A, Supplementary Table 1,2**).

**Table 1.** Colocalization analyses of COVID-19 outcomes and other diseases at the identified sQTL loci.

| sQTL locus (rsID) | chr | pos (b38) | EA | NEA | intron junction | COVID-19 outcome | Other outcome | PP* |
| --- | --- | --- | --- | --- | --- | --- | --- | --- |
| *ATP11A*<br>(rs12585036) | 13 | 112881427 | C | T | chr13:112,875,941-112,880,546 ↑<br>(lung, WBC) | critical illness ↓ | idiopathic pulmonary fibrosis ↑<br>[PMID: 31710517] | 1 |
| *DPP9*<br>(rs12610495) | 19 | 4717660 | A | G | chr19:4,814,337-4,717,615 ↑<br>(lung) | critical illness ↓ | idiopathic pulmonary fibrosis ↓<br>[PMID: 31710517] | 1 |
| *NPNT*<br>(rs34712979) | 4 | 105897896 | A | G | chr4:105,898,001-105,927,336 ↑<br>(lung) | critical illness ↓ | FEV1/FVC ratio ↓<br>[PMID: 30804560] | 1 |
|  |  |  |  |  |  |  | COPD ↑<br>[PMID: 30804561] | - |
|  |  |  |  |  |  |  | Asthma ↑<br>[PMID: 31959851] | - |
| *OAS1*<br>(rs10774671) | 11 | 112919388 | G | A | chr12:112,917,700-112,919,389 ↑<br>(lung) | critical illness ↓ | systemic lupus erythematosus ↓<br>[PMID: 33272962] | - |
|  |  |  |  |  |  |  | chronic lymphocytic leukemia ↑<br>[PMID: 28165464] | - |
| *MUC1*<br>(rs4072037) | 1 | 155192276 | C | T | chr1:155,192,310:155,192,786 ↑<br>(lung) | reported infection ↑ | inflammatory bowel disease ↓<br>[PMID: 28067908] | 1 |
|  |  |  |  |  |  |  | gastric cancer ↓<br>[PMID: 26098866] | - |
|  |  |  |  |  |  |  | gout ↓<br>[PMID: 33832965] | - |
|  |  |  |  |  |  |  | Urate level ↓<br>[PMID:33462484] | 1 |

EA: effect allele, NEA: non-effect allele, PP: a posterior probability that there is an association for both gene expression/splicing and COVID-19 outcomes,
which is driven by the same causal variant. PP was only estimated when there is an available GWAS summary statistics of European ancestry.

**Fig. 2.**
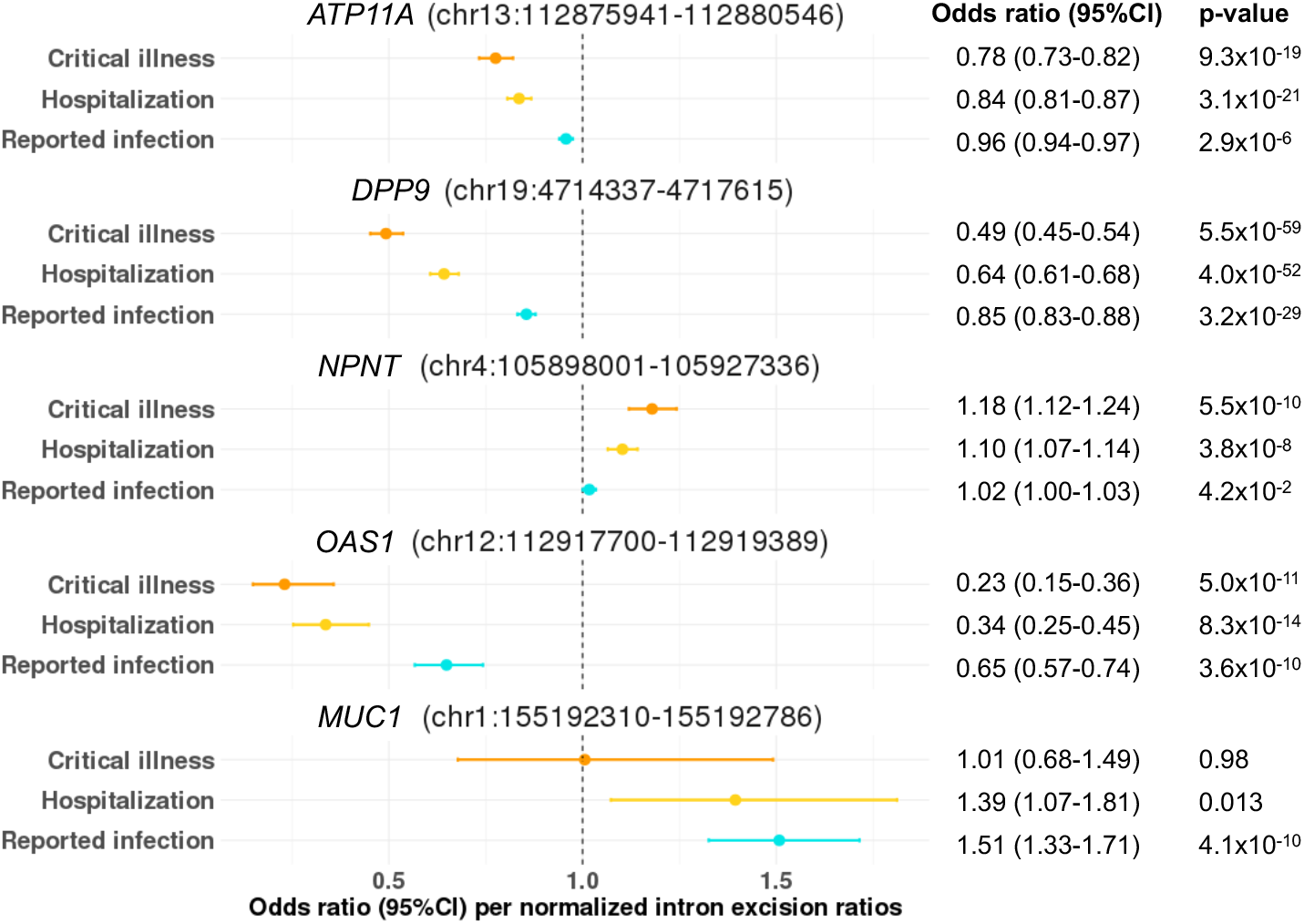
MR estimates of the effect of gene splicing at *ATP11A, DPP9, NPNT, OAS1*, and *MUC1* with COVID-19 outcomes. Forest plot showing odds ratio and 95% confidence interval from two sample Mendelian Randomization analyses (two-sided). P values are unadjusted. Unit: standard deviation of intron excision ratios as quantified by LeafCutter.

We additionally identified that *ATP11A* sQTL, rs12585036:T>C, which increases the excision of the intron junction at chr13:112,875,941-112,880,546 in lung, was associated with protection against all three adverse COVID-19 outcomes with odds ratio [OR] per normalized intron excision ratio [the proportion of reads supporting each alternatively excised intron identified by LeafCutter^20^] of 0.78 (95%CI: 0.73-0.82, p=9.3×10^−19^) for critical illness, OR of 0.84 (95%CI: 0.81-0.87, p=3.1×10^−21^) for hospitalization, and OR of 0.96 (0.94-0.97, p=2.9×10^−6^) for reported infection (**Fig. 2, Extended Data Fig. 1B, Supplementary Table 1**).

We also found novel associations of *DPP9* sQTL, rs12610495:G>A, which increases the excision of the intron junction at chr19:4,714,337-4,717,615 in lung, with protection against all three adverse COVID-19 outcomes (OR: 0.49, 0.45-0.54, p=5.5×10^−59^ for critical illness, OR: 0.64, 0.61-0.68, p=4.0×10^−52^ for hospitalization, and OR: 0.85, 0.83-0.88, p=3.2×10^−29^ for reported infection, **Fig. 2**). DPP9 has two major isoforms, DPP9L (longer isoform, 863 aa, ENST00000262960) and DPP9S (shorter isoform, 863 aa, ENST00000598800)^25^, and they differ in the presence of the skipping of exon 9 (chr19:4,715,529-4,716,152), which overlaps the intron junction at chr19:4,714,337-4,717,615 (**Extended Data Fig. 1C, Supplementary Table 1**).

*NPNT* rs34712979:A>G, which increases the intron junction of chr4:105,898,001-105,927,336 (**Extended Data Fig. 1D**), was associated with increased risk of severe COVID-19 outcomes (critical illness and hospitalization) but with marginal evidence of increased risk of SARS-CoV-2 infection (OR: 1.18, 1.12-1.24, p=5.5×10^−10^ for critical illness, OR: 1.10, 1.07-1.14, p=3.8×10^−8^ for hospitalization, and OR: 1.02, 1.00-1.03, p=4.2×10^−2^ for reported infection, **Fig. 2, Supplementary Table 1**). The *NPNT* sQTL, rs34712979-A allele, creates a NAGNAG splice acceptor site, which results in additional in-frame AGT codon, coding for serine, at the 5’ splice site of exon 2^26^.

Gene splicing in the two genes in the gene cluster located in chr1 q21.3-q22, namely *MUC1* and *THBS3*, in lung were also associated with reported SARS-CoV-2 infection, but not with COVID-19 severity phenotypes (**Fig. 2, Supplementary Table 1**). Given that the *MUC1* and *THBS3* sQTL SNPs are in high LD with each other (rs4072037:C>T and rs2066981:A>G; r2=0.98 in European population of 1000G), it was challenging to distinguish which genes were more likely to be causal between *MUC1* and *THBS3*. Nevertheless, *MUC1* sQTL SNP, rs4072037:C>T, is a recognized splice variant which influences the 3’ splice site selection of exon2^27^. In GTEx, the rs4072037-T allele was associated with decreased levels of the intron junction of chr1:155,192,310:155,192,786 (**Extended Data Fig. 1E**), which leads to transcripts that have an alternative 27 bp intron retention event at the start of exon 2^27^. We demonstrated the rs4072037-C allele, which increases the levels of the intron junction of chr1:155,192,310:155,192,786, was associated with increased risk of reported SARS-CoV-2 infection. Given the well-known evidence of the splicing event of *MUC1*, it is more likely that *MUC1* gene splicing has an impact of SARS-CoV-2 infection.

All of the above associations of gene splicing, except for *MUC1*, were more pronounced with more severe outcomes (**Fig. 2**). We also applied MR for another hospitalization phenotype compared within COVID-19 cases (“hospitalization vs non-hospitalization amongst individuals with laboratory-confirmed SARS-CoV-2 infection”, which corresponds to B1 phenotype in COVID-19 HGI^11^). We confirmed the associations with gene splicing in *ATP11A* and *DPP9* (*ATP11A*: OR 0.87, 0.81-0.94, p=4.6×10^−4^, and *DPP9*: OR 0.83, 0.73-0.94, p=4.9×10^−3^) but not with gene splicing in *NPNT* and *OAS1* (**Supplementary Table 3**).

### Colocalization and European-only MR analyses

To test whether confounding due to linkage disequilibrium may have influenced the MR estimates, we tested the probability that alternative splicing and the COVID-19 outcomes shared a single causal signal using colocalization analyses, as implemented in coloc^28^. These colocalization analyses were performed using data from individuals of European ancestry, as this method assumes that the two GWASs tested were derived from the individuals with the similar LD structure. We next performed MR as a sensitivity analysis by using the data only from individuals of European descent to ensure that the MR results were not biased from population stratification^29^.

Amongst the MR-prioritized splicing events (29 genes in lung and 20 genes in whole blood, **Supplementary Table 1,2**), we selected the splicing events with high (>0.8) posterior probability for hypothesis 4 (PP, that there is an association for both gene expression/splicing and COVID-19 outcomes, which is driven by the same causal variant), and with a support from European-descent only MR analysis (p<5.1×10^−6^, where the p-value threshold was adopted from the main analysis) (**Fig1, 3, Extended Data Fig. 1**). These analyses retained 8 genes (*ABO, ATP11A, DPP9, GBAP1, MUC1, NPNT, OAS1, THBS3*) whose alternative splicing in lung influenced at least one COVID-19 outcome and 2 genes (*MOSPD3, OAS1*) whose alternative splicing in whole blood influenced at least one COVID-19 outcome (**Supplementary Table 4,5**).

A posterior probability (PP) that alternative splicing of *ATP11A* in lungs and COVID-19 outcomes shared a single causal signal in the 1Mb locus around the *cis*-sQTL was 1.00 for critical illness, 1.00 for hospitalization due to COVID-19, and 0.98 for reported infection (**Fig. 3A**). Alternative splicing of *DPP9* had as high PP as 0.96 for critical illness, 0.96 for hospitalization, and 0.95 for reported infection (**Fig. 3B**). Alternative splicing of *NPNT* had as high PP as 1.00 for critical illness, and 1.00 for hospitalization, but had as low PP as 0.02 for reported infection (**Fig. 3C**). All of these gene splicing had a support from European-descent only MR analysis with similar effect estimates as the main analyses (p<5.1×10^−6^, **Supplementary Table 4,5**).

**Fig. 3.**
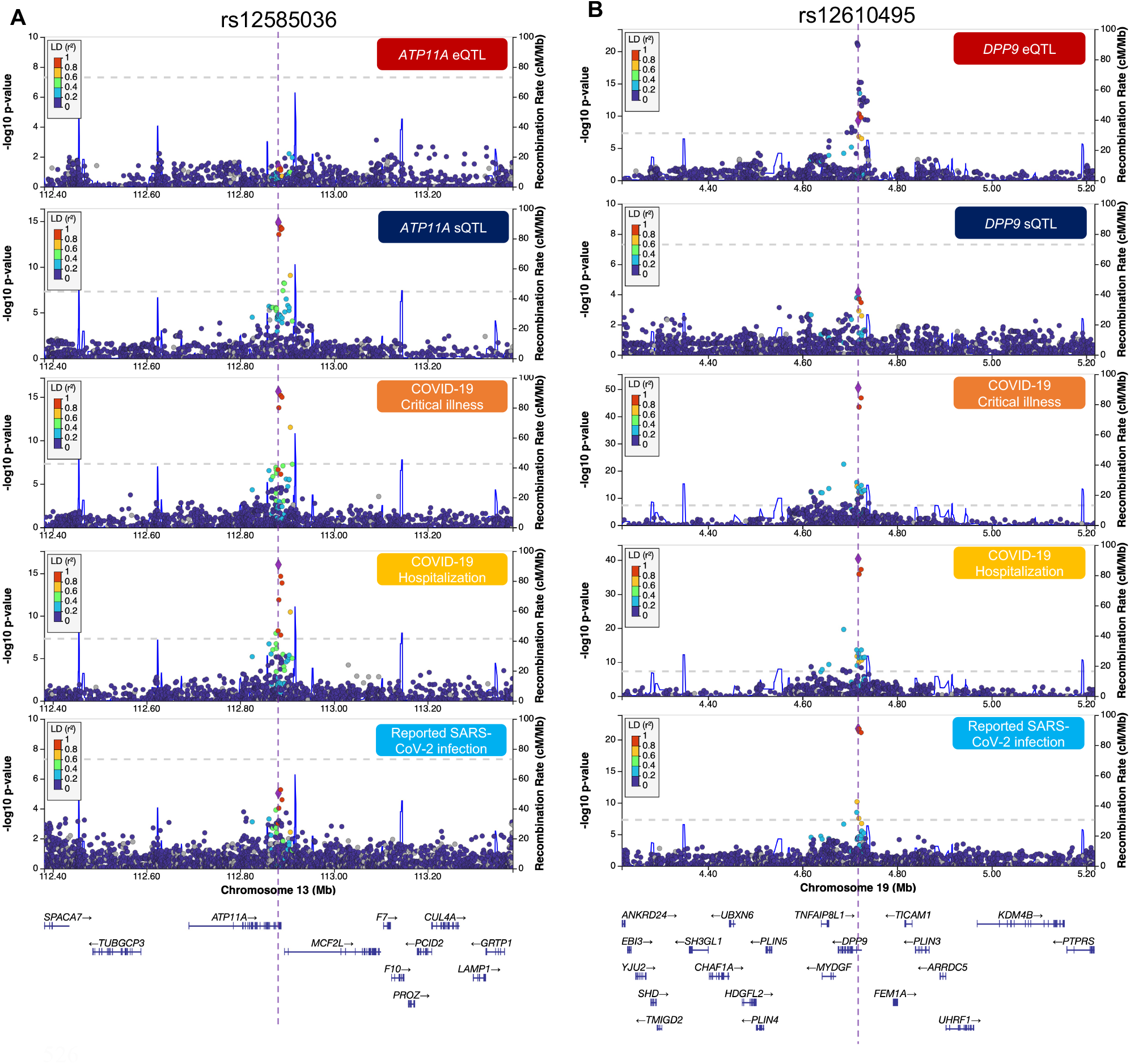
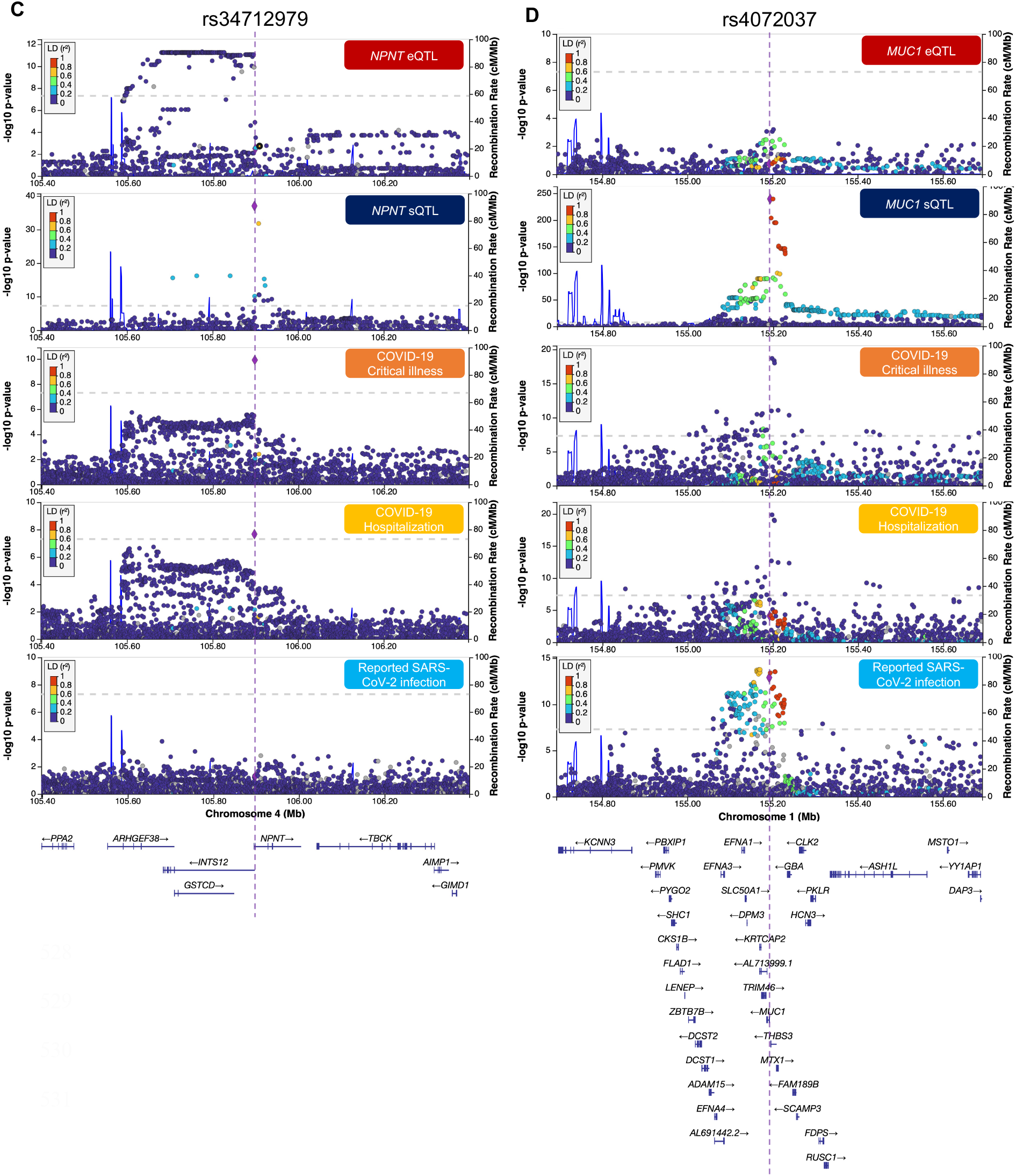
LocusZoom plots demonstrating the colocalization of the genetic determinants of mRNA expression, gene splicing levels and COVID-19 outcomes. LocusZoom plots of genetic signal of 1MB region around each *cis*-sQTL and COVID-19 outcomes, color shows SNPs in the region in LD (r^2^) to the *cis*-sQTL. (A) *ATP11A* (B) *DPP9* (C) *NPNT* (D) *MUC1*

### Colocalization with eQTL GWASs from GTEx

Next, to understand if total gene expression levels in lung and whole blood were associated with COVID-19 outcomes, without respect to the splicing or isoforms, we performed MR and colocalization analyses using expression quantitative trait loci (eQTLs) in lung and whole blood from GTEx. The total expression of *GBAP1* had MR evidence with reported SARS-CoV-2 infection (p=5.9×10^−8^) and the eQTLs of *ABO, GBAP1*, and *MOSPD3*, had high colocalization with COVID-19 outcomes (PP >0.8, **Supplementary Table 6,7**). Thus, for those genes, we could not clarify whether the associations with COVID-19 outcomes were driven by either total gene expression or the spliced isoform expression or both.

### Influence of prioritized sQTLs on other diseases

We next searched for the effects of the sQTLs, which influence COVID-19 outcomes, on other diseases. To do so, we used the Open Targets Genetics^30,31^ (https://genetics.opentargets.org) and identified GWAS lead variants that were in LD (r^2^>0.80) with COVID-19 associated sQTLs. We found that the *DPP9* sQTL (rs12610495:G>A) was associated with decreased risk of idiopathic pulmonary fibrosis (IPF)^32^ and the same allele confers the protection against COVID-19 severity. Interestingly, we found the opposite direction of effects between IPF and COVID-19 in the sQTL for *ATP11A*, where the rs12585036-C allele was associated with increasing risk of IPF and decreased risk for COVID-19 severity. The *NPNT* sQTL had similar trend that rs34712979-A allele, which was protective for COVID-19 severity, was associated with increased risk of COPD (*i*.*e*. lower FEV1/FVC ratio, a spirometry measurement used to diagnose COPD)^33,34^ and with increased risk of asthma^35^. The *OAS1* sQTL (rs10774671:A>G) was associated with a decreased risk of systemic lupus erythematosus (in the East Asian population)^36^ and with an increased risk of chronic lymphocytic leukemia^37^. The *MUC1* sQTL (rs4072037:T>C) was associated with decreased risk of gastric cancer^38^, gout^39^, and inflammatory bowel disease^40^.

We then performed colocalization analyses in each sQTL locus between COVID-19 outcomes and the other associated diseases/phenotypes if the effects were supported in European population and there was genome-wide GWAS summary data available. IPF^32^ and critical illness due to COVID-19 were highly colocalized at the *ATP11A* and *DPP9* sQTL loci with PP of 1.00. The FEV1/FVC ratio also colocalized with critical illness due to COVID-19 with a PP of 1.00. Inflammatory bowel disease^40^ and urate level (which is the cause of gout) also colocalized with reported SARS-CoV-2 infections at the *MUC1* locus (both PPs are 1). We found no GWAS summary statistics available for the other diseases (**Table 1**). These lines of evidence suggest that IPF and COVID-19 severity, COPD/asthma and COVID-19 severity, and IBD and COVID-19 susceptibility may share causal genetic determinants in these loci, respectively. The full results and the data used are summarized in **Table 1**.

### The tissue and cell-type specific expression of the associated genes

To assess the relevant tissues and cell-types for the associated gene splicing – COVID-19 outcome relationships, we evaluated the associated gene expression in lung and peripheral blood mononuclear cell (PBMC) of healthy controls, as well as in lung of COVID-19 patients. In the consensus transcript expression levels from Human Protein Atlas (HPA)^41^ and GTEx, the expression of *ATP11A*, and *NPNT* were highly enriched in normal lung tissue (**Fig. 4A**). At a single-cell resolution, in normal lung, *MUC1*,

**Figure 4.**
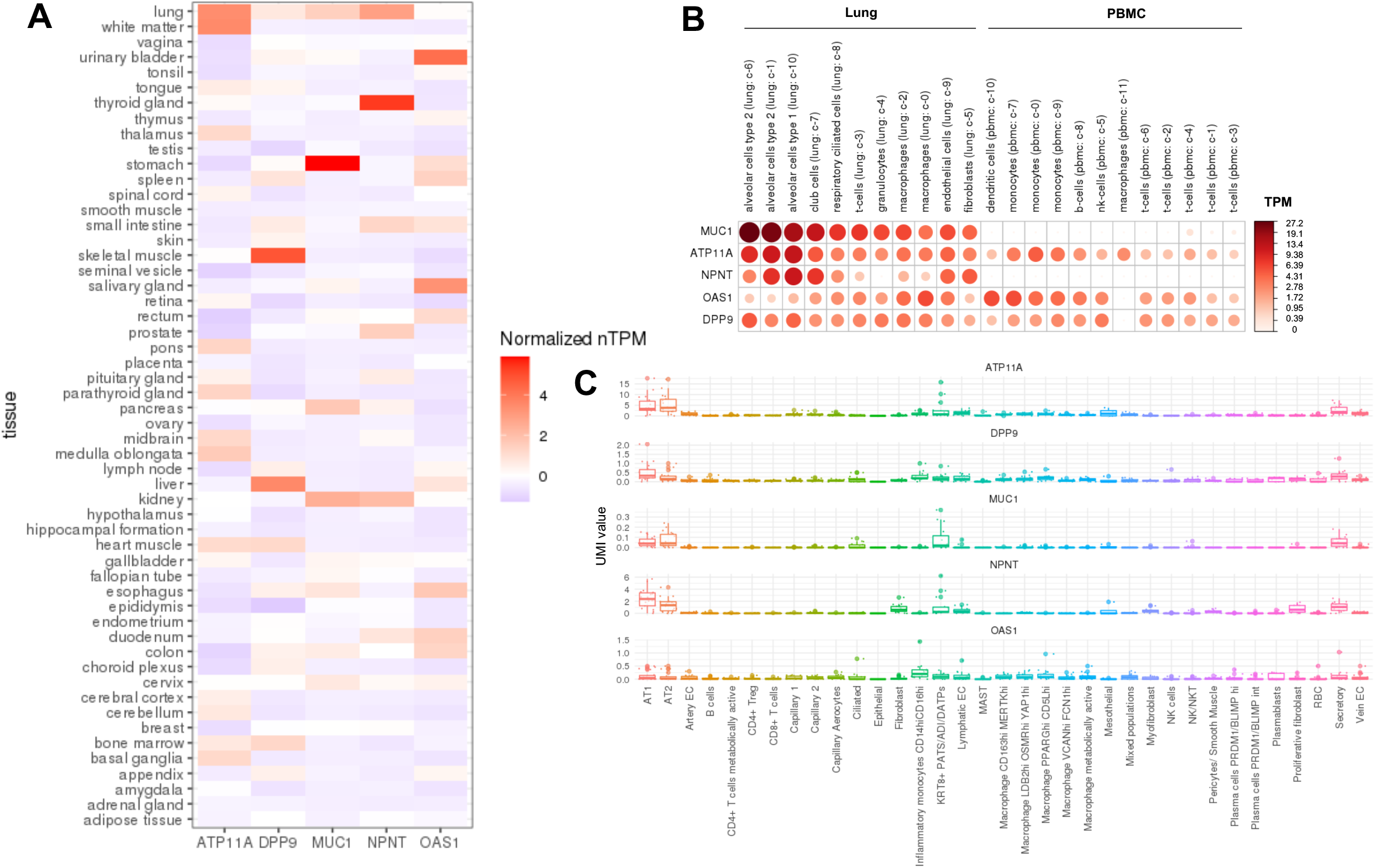
The gene expression in lung and peripheral blood mononuclear cell (PBMC). A. The consensus transcript expression levels summarized per gene in 54 tissues in Human Protein Atlas (HPA), which was calculated as the maximum transcripts per million value (TPM) value for each gene in all sub-tissues categories of each tissue, based on transcriptomics data from HPA and GTEx. B. RNA single cell type tissue cluster data (transcript expression levels summarized per gene and cluster) of lung (GSE130148) and peripheral blood mononuclear cell (PBMC) (GSE112845) were visualized using log_10_(protein-transcripts per million [pTPM]) values. Each c-X annotation is taken from the clustering results performed in Human Protein Atlas. C. Single-cell RNA expression profile of 23 lung COVID-19 autopsy donor tissue samples from GSE171668^43^. The mean value of RNA expression of the cells annotated in the same subcategory was represented as a dot per each sample. The cell type annotation was manually performed in the original publication^43^. AT1: alveolar type 1 epithelial cells, AT2: alveolar type 2 epithelial cells, EC: endothelial cells, *KRT8*^+^ PATS/ADI/DATPs: *KRT8*^+^ pre-alveolar type 1 transitional cell state, MAST: mast cells, XXhi: XX (gene expression) ^*high*^ cells, RBC: red blood cells.

*ATP11A* and *NPNT* were specifically enriched in alveolar type 1 and 2 epithelial cells (**Fig. 4B**), which are known to play important roles in regeneration of alveolar epithelium following lung injury^42^. In 23 lung COVID-19 autopsy donor tissue samples from GSE171668^43^, the expression of *ATP11A, DPP9, MUC1*, and *NPNT* were also enriched in alveolar type 1 and 2 epithelial cells (**Fig. 4C**). Taken together, these expression evidence suggests that the gene splicing – COVID-19 outcome relationships for *ATP11A, DPP9, MUC1*, and *NPNT* were highly likely to be relevant in lung tissue, especially alveolar epithelial cells for *ATP11A, MUC1*, and *NPNT*.

## Discussion

Despite current vaccines and therapeutic options, hospitalization for COVID-19 remains high in many countries. Thus, there remains a need for additional therapies, which in turn, requires target identification and validation^44^. In this large-scale two-sample MR study of 8,172 gene splicing events assessed for their effect upon three COVID-19 outcomes in up to 158,840 COVID-19 cases and 2,782,977 population controls, we provide evidence that alternative splicing of *OAS1, ATP11A, DPP9* and *NPNT* in lung influences COVID-19 severity, and alternative splicing of *MUC1* in lung influences COVID-19 susceptibility. Moreover, these genetic mechanisms are likely shared with other diseases, such as *ATP11A* and *DPP9* with IPF, suggesting the potential to be targeted for multiple diseases. Our results provide rationale to target gene splicing in lung as COVID-19 treatment by means of therapeutic modalities such as splice-switching oligonucleotides (SSOs).

SSOs are a type of antisense oligonucleotides which are generally 15–30 nucleotides in length and are designed as complementary to specific regions of mRNA with increased stability against endogenous nucleases due to chemical modifications^45^. SSOs can prevent splicing promoting factors from binding to the target pre-mRNA, which can modulate alternative splicing^46^. Some SSOs have been already approved by FDA, such as eteplirsen for Duchenne’s muscular dystrophy, which induces the skipping of exon 51 of *DMD* gene, and nusinersen for spinal muscle atrophy, which induces the expression of exon 7 of *SMN2* gene^45^. Although delivery to target tissues after systemic administration has been a key challenge in the development of SSO drugs^45,47^, lung might be advantageous tissue to target since it is possible to deliver drugs directly to lung through inhalation and/or bronchoscopy.

There are multiple sources of biological evidence which might support the relevance of *ATP11A* in COVID-19. *ATP11A* is a member of the P4-ATPase family, which codes for a phospholipid flippase at the plasma membrane and translocates phosphatidylserine (PtdSer) from the outer to the inner leaflet of plasma membranes. Cells that undergo apoptosis, necroptosis or pyroptosis expose PtdSer on their surface through P4-ATPase, of which *ATP11A* is cleaved by caspase during apoptosis^48^. While less functionally defined, given the high expression of *ATP11A* in the alveolar type 2 epithelial cells, *ATP11A* sQTL (rs12585036:C>T) may impact the cell death of alveolar type 2 epithelial cells, the resident stem cell population in lung^49^, and may impair regeneration of alveolar epithelium following lung injury.

Our MR analysis showed that the excision of intron junction chr19:4,814,337-4,717,615, which may be preferentially spliced in DPP9L than in DPP9S, was associated with reduced risk of COVID-19 severity. While both DPP9L and DPP9S can be active^25^, DPP9S is present in the cytosol, and DPP9L localizes preferentially to the nucleus, which may be attributed to a classical monopartite nuclear localization signal (RKVKKLRL) in the N-terminal extension of DPP9L^25^. Although it is not fully understood how the *DPP9* sQTL (rs12610495:A>G) regulates alternative splicing of *DPP9*, rs12610495-G allele creates a GGGG motif (from GGAG sequence), which may have an effect on alternative splicing of some genes^50^. DPP9 interacts with NLRP1 and represses inflammasome activation^51^ and pyroptosis, which are now recognized as important mechanisms interrupting the viral replication cycle and preventing viral amplification of SARS-CoV-2^52^. Moreover, targeting inflammasome-mediated hyperinflammation in COVID-19 patients may also prevent chronic phase of COVID-19 pathophysiology in vivo^52^.

Our MR analyses supported that the *MUC1* gene splicing associated with susceptibility of SARS-CoV-2 infection. *MUC1* is a mucin, which is also called KL-6 in human, and the serum level of KL-6 is used as a biomarker for interstitial lung diseases^53^. rs4072037-T allele was associated with the decreased level of the intron junction of chr1:155,192,310:155,192,786, which corresponds to transcripts with an alternative 27 bp intron retention event at the start of exon 2^27^. *MUC1* exon 2 harbors a variable number of tandem repeats (VNTR) that contains 20 to 125 repeats of a 60 bp coding sequence that determines the length of a heavily glycosylated extracellular domain^54^. Although rs4072037 may or may not control the alternative splicing of VNTR region^38^, in addition to the alternative 27 bp intron retention^27^, the VNTR length of *MUC1* is associated with several renal phenotypes in recent study in UK Biobank^55^. The rs4072037-T allele has been shown to be associated with increased risk of gastric cancer in both European and East Asian ancestries^38,56^ and it is hypothesized that the *MUC1* gene is involved in response to *H*.*pylori* colonization in gastric epithelial surfaces. Taken together, it is plausible that alternative splicing of *MUC1* in lung alveolar cells has an impact on SARS-CoV-2 infection.

This study has limitations. First, the excised intron junction was quantified in an annotation-free manner using LeafCutter^20^, without respect to the level of transcripts nor isoforms. It is thus important for future work to map those disease-relevant alternative splicing events to the corresponding isoform or protein product, by means of emerging technologies such as long-read sequencing^57^ and high-throughput protein quantification^58^. We anticipate our findings, and others, will motivate the ongoing effort to do so. Second, we used MR to test the effect of gene splicing measured in a non-infected state since the effect of the *cis-*sQTLs upon gene splicing was estimated in individuals who had not been exposed to SARS-CoV-2. Given the dynamic gene regulation of splicing during infection^59^, gene splicing could be altered once a person experiences SARS-CoV-2 infection. Thus, the MR results presented in this paper should be interpreted as an estimation of the effect of gene splicing during uninfected state. Future studies may help to clarify if the same *cis-*sQTLs regulate alternative splicing during infection. Third, it was not our goal to identify all alternative splicing that affects COVID-19 outcomes, but rather to provide strong evidence for a small set of genes with strong MR and colocalization evidence. Thus, we acknowledge a high false-negative rate of our study design.

Although we attempted to include the data from individuals of diverse ancestries, most individuals were of European descent. Therefore, we could not confirm that our findings could be robustly transferrable to individuals of other populations. Fourth, while our findings demonstrate that effects of alternative splicing in lung and whole blood since these are relevant to COVID-19 severity, we recognize that these splicing events may not be unique to these tissues and thus the estimated effects of splicing may represent the action of these same alternative transcripts in other tissues. Lastly, a recent paper^60^ reported that the shared genetic signal between IPF and COVID-19 outcomes at the *DPP9* and *ATP11A* loci are likely driven by the difference of total expression in whole blood, which was supported by colocalization using eQTLGen^61^ dataset. However, eQTLGen consists of multi-ancestry cohorts and could be affected by the bias due to LD. Moreover, we demonstrated that both *DPP9* and *ATP11A* expression were enriched in alveolar cells in lung, compared to whole blood. Thus, we concluded that it is more likely that the alternative splicing of *DPP9* and *ATP11A* in lung is important both for IPF and COVID-19 severity.

In conclusion, we have used genetic determinants of gene splicing and COVID-19 outcomes obtained from large-scale studies and found compelling evidence that splicing events in *OAS1, ATP11A, DPP9, NPNT*, and *MUC1* have causal effects on COVID-19 severity and susceptibility. Interestingly, the available evidence suggests shared genetic mechanisms for COVID-19 severity with IPF at *ATP11A* and *DPP9* loci, and with chronic obstructive lung diseases at the *NPNT* locus. Taken together, our study highlights the importance of alternative splicing both in COVID-19 and other diseases, which could be further investigated for drug discovery such as SSOs.

## Methods

### Splicing quantitative trait loci (sQTL) GWASs

We obtained all conditionally independent splicing quantitative trait loci (sQTLs) in lung (N=515, of which 452 were of European ancestry) and whole blood (N=670, of which 570 were of European ancestry) that act in *cis* (in a +/- 1Mb window around the transcription start site of each gene with normalized intron excision ratios from LeafCutter^20^ (5% FDR per tissue). Conditional independent sQTLs were mapped using stepwise regression in GTEx consortium^19^. Intron excision ratios are the proportion of reads supporting each alternatively excised intron identified by LeafCutter^20^. We selected lung and whole blood since they are the two major tissues relevant to COVID-19 pathophysiology.

### COVID-19 GWASs

To assess the association of *cis-*sQTLs with COVID-19 outcomes, we used COVID-19 GWAS meta-analyses results from the COVID-19 Host Genetics Initiative (HGI) release 7^11^ (https://www.covid19hg.org/results/r7/). The outcomes tested were critical illness, hospitalization, and reported SARS-CoV-2 infection (named A2, B2, and C2, respectively by the COVID-19 HGI).

Critically ill COVID-19 cases were defined as those individuals who were hospitalized with laboratory-confirmed SARS-CoV-2 infection and who required respiratory support (invasive ventilation, continuous positive airway pressure, Bilevel Positive Airway Pressure, or continuous external negative pressure, high-flow nasal or face-mask oxygen) or who died due to the disease. Simple supplementary oxygen (e.g. 2 liters/minute via nasal cannula) did not qualify for case status. Hospitalized COVID-19 cases were defined as individuals hospitalized with laboratory-confirmed SARS-CoV-2 infection (using the same microbiology methods as for the critically ill phenotype), where hospitalization was due to COVID-19 related symptoms. Reported SARS-CoV-2 infection was defined as laboratory-confirmed SARS-CoV-2 infection or electronic health record, ICD coding or clinically confirmed COVID-19, or self-reported COVID-19 (for example, by questionnaire), with or without symptoms of any severity. Controls were defined in the same way across all three outcomes above as everybody that is not a case—for example, population controls.

In a sensitivity analysis, we also used another hospitalization phenotype (named B1 in the COVID-19 HGI), where cases were hospitalized COVID-19 cases and controls were defined as non-hospitalized individuals with laboratory-confirmed SARS-CoV-2 infection.

### Two-sample Mendelian randomization

We used two-sample mendelian randomization (MR) analyses using “TwoSampleMR v0.5.6” R package^62^ to screen and test the potential role of alternative splicing in lung and whole blood to influence COVID-19 outcomes. In two-sample MR, the effect of SNPs on the exposure and outcome are taken from separate GWASs. Two-sample MR often improves statistical power and is less biased than one-sample MR, with potential bias in the direction to the null^63^. MR is less affected by confounding and reverse causality than observational epidemiology studies. The MR framework is based on three main assumptions: First, the SNPs are robustly associated with the exposure (*i*.*e*. a lack of weak instrument bias). We validated that all *cis*-sQTLs had a F-statistic >10, corresponding to T-statistics >3.16, which indicate a low risk of weak instrument bias in MR analyses^64^. Second, the SNPs are not associated with any confounding factors for the relationship between the exposure and the outcome. Third, the SNPs have no effect on the outcome that is independent of the exposure (*i*.*e*. a lack of horizontal pleiotropy), which is the most challenging assumption to assess. Nevertheless, in order to reduce the risk of horizontal pleiotropy, we selected *cis-*sQTLs as instrumental variables, as *cis-*SNPs that reside close to the genes are more likely to have an effect on the outcomes by directly influencing the splicing of the gene. Palindromic *cis*-sQTLs with minor allele frequencies (MAF) >0.42 were removed prior to MR to prevent allele-mismatches. For gene splicing with a single (sentinel) *cis-*sQTL, we used the Wald ratio to estimate the effect of each splicing event on each of the three COVID-19 outcomes. For any gene splicing event with multiple conditionally independent *cis*-sQTLs, an inverse variance weighted (IVW) method was used to meta-analyze their combined effects. After harmonizing the *cis-*sQTLs with COVID-19 GWASs, a total of 5,999 splicing events in 4,477 genes (6,110 matched *cis-*sQTLs) in lung and a total of 3,769 splicing events in 2,779 genes (3,883 matched *cis-*sQTLs) in whole blood were used for the MR analyses across the three COVID-19 outcomes. We applied the Bonferroni corrected p-value (5.1×10^−6^) to adjust for multiple testing, by using the number of splicing events tested (5,999 in lung and 3,769 in whole blood, in total 8,172).

### European-only MR analyses and colocalization analysis

Next, we assessed potential confounding by population stratification and linkage disequilibrium (LD). To do so, we first repeated MR analyses using the data from individuals of European ancestry. We obtained the effect estimate of each *cis-*sQTL from the data mapped in European-American subjects. If the *cis-* sQTL was missing in the summary data mapped in European-ancestry subjects, we removed those *cis-* sQTLs in the analyses. The three COVID-19 outcomes GWASs results were also replaced by the meta-analyses summary results of European-ancestry subjects.

We also evaluated whether the gene splicing and COVID-19 outcomes shared a common etiological genetic signal and that the MR results were not biased by linkage disequilibrium (LD) using colocalization analyses with the data from individuals of European ancestry. Specifically, for each of these MR significant splicing events, a stringent Bayesian analysis was implemented in “coloc v 5.1.0.1” R package to analyze all variants in 1MB genomic locus centered on the *cis*-sQTL. Colocalizations with posterior probability for high colocalization (PP >0.8), that is, that there is an association for both gene splicing and COVID-19 outcomes, and they are driven by the same causal variant were considered to colocalize, which means that the exposure and the outcome shared a single causal variant.

### Mendelian randomization and Colocalization with eQTL GWASs from GTEx

To understand if the total gene expression levels in lung and whole blood were associated with COVID-19 outcomes, without respect to the splicing or isoforms, we similarly performed MR and colocalization analyses using expression quantitative trait loci (eQTLs) in lung and whole blood from GTEx v.8^19^ (lung: N=515, of which 452 were of European ancestry, and whole blood: N=670, of which 570 were of European ancestry) by restricting the regions within 1 Mb of each QTL. The genetic instruments were conditionally independent eQTLs for the prioritized sQTL genes in lung and/or whole blood, all of which had strong support for colocalization between sQTLs and COVID-19 outcomes.

### Influence of identified sQTLs on other diseases

We assessed the effects of the sQTLs which influence COVID-19 outcomes on other diseases. Pleiotropic search was performed using Open Targets Genetics^30,31^ (https://genetics.opentargets.org) and identified any GWAS lead variants that are in LD (r^2^>0.80) with those sQTLs. Colocalization analyses were performed using “coloc v 5.1.0.1” R package when there is available GWAS summary statistics of European ancestry.

### The tissue and cell-type specific expression of the associated genes

To assess the relevant tissues and cell-types for the associated gene splicing – COVID-19 outcome relationships, we evaluated the associated gene expression in lung and peripheral blood mononuclear cell (PBMC) of healthy controls, as well as in lung of COVID-19 patients. We first downloaded consensus transcript expression levels summarized per gene in 54 tissues in Human Protein Atlas (HPA)^41^, which was calculated as the maximum transcripts per million value (TPM) value for each gene in all sub-tissues categories of each tissue, based on transcriptomics data from HPA and GTEx. We also downloaded the single-cell type transcriptomic analyses, where we used all cell types in lung (GSE130148^65^) and peripheral blood mononuclear cell (PBMC) (GSE112845^66^). We visualized RNA single cell type tissue cluster data (transcript expression levels summarized per gene and cluster), using log_10_(protein-transcripts per million [pTPM]) values with “corrplot v 0.92” R package. Lastly, we obtained single-cell RNA expression profile of 23 lung COVID-19 autopsy donor tissue samples from GSE171668^43^. We calculated “pseudo-bulk” RNA expression per each cell type by taking the mean value of RNA expression of the cells annotated in the same subcategory.

## Data Availability

All data produced are available online at
https://www.covid19hg.org/results/r7/
https://www.gtexportal.org/home/datasets
https://github.com/genomicsITER/PFgenetics
https://www.ebi.ac.uk/gwas/
https://www.proteinatlas.org/about/download
https://www.ncbi.nlm.nih.gov/geo/query/acc.cgi?acc=GSE171668

## Data availability

Data from GTEx consortium (GTEx project v8^19^) are available from the referenced peer-reviewed studies or their corresponding authors, as applicable. Summary statistics for eQTLs and sQTLs from GTEx v8 are publicly available for download on the GTEx website (gtexportal.org/home/datasets). Summary statistics for the COVID-19 outcomes are publicly available for download on the COVID-19 Host Genetics Initiative website^67^ (www.covid19hg.org). The consensus transcript expression levels and RNA single cell type tissue cluster data were downloadable from the Human Protein Atlas^41^ website (www.proteinatlas.org/about/download). The processed single-cell RNA expression profile of 23 lung COVID-19 autopsy donor tissue sample are freely available in the Gene Expression Omnibus (GEO, https://www.ncbi.nlm.nih.gov/geo/) under accession number GSE171668^43^.

## Code availability

All code for data management and analysis is archived online at github.com/richardslab/COVID19-sQTLMR for review and reuse.

---

**Extended Data Fig. 1.**
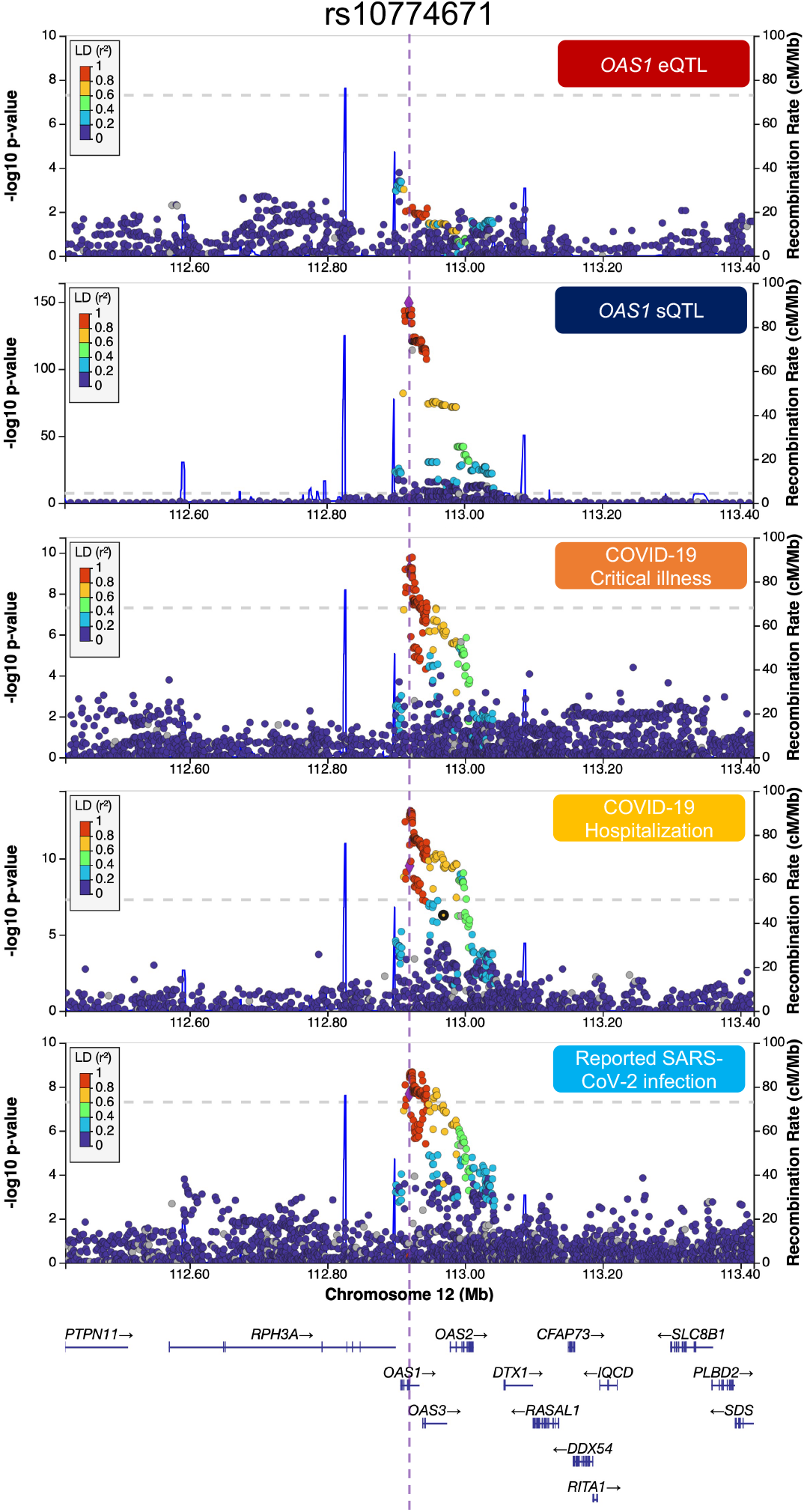
LocusZoom plots demonstrating the colocalization of the genetic determinants of mRNA expression, gene splicing levels for *OAS1* and COVID-19 outcomes. LocusZoom plots of genetic signal of 1MB region around each *cis*-sQTL and COVID-19 outcomes, color shows SNPs in the region in LD (r^2^) to the *cis*-sQTL for *OAS1*.

**Extended Data Fig. 2.**
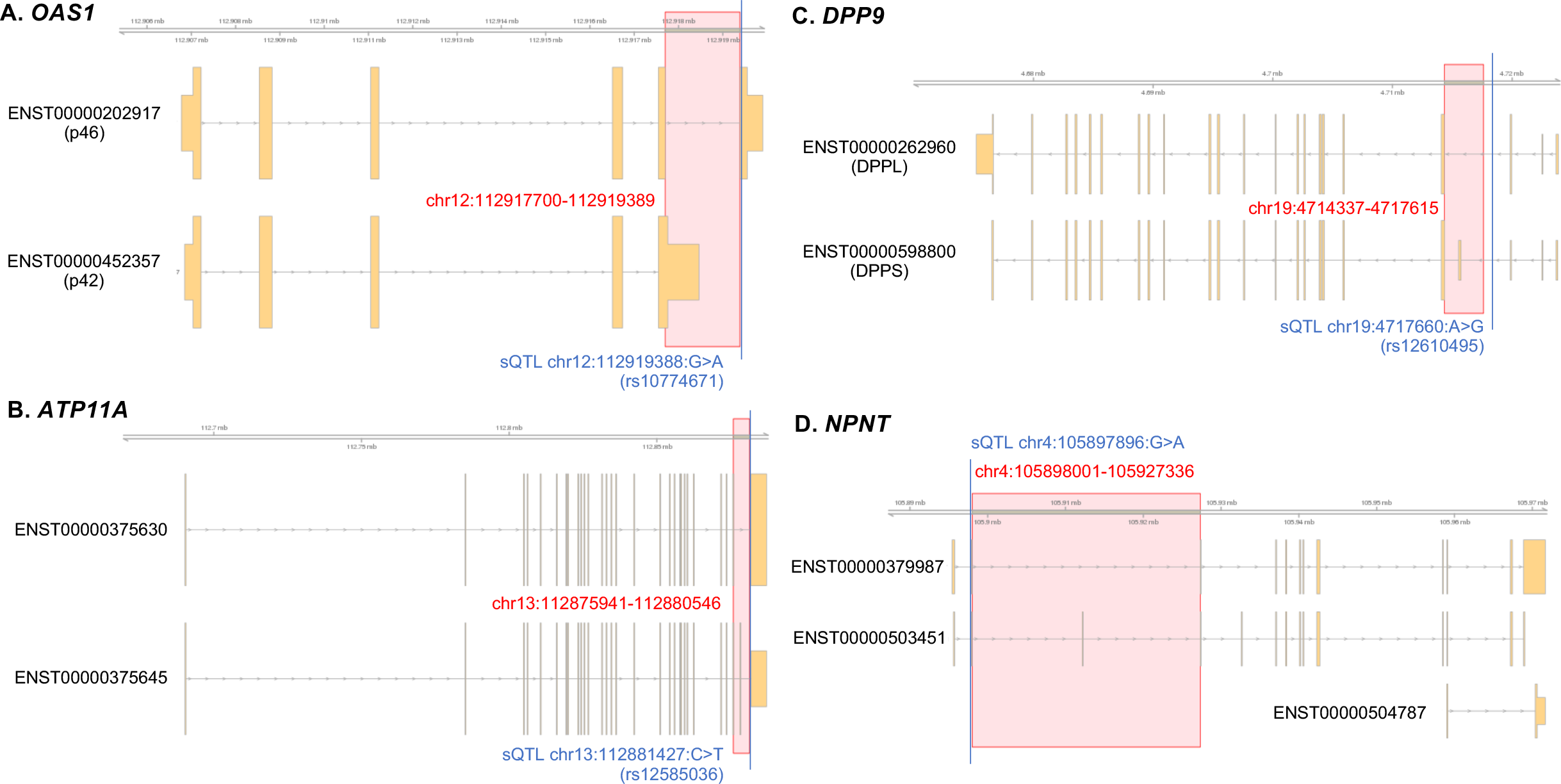
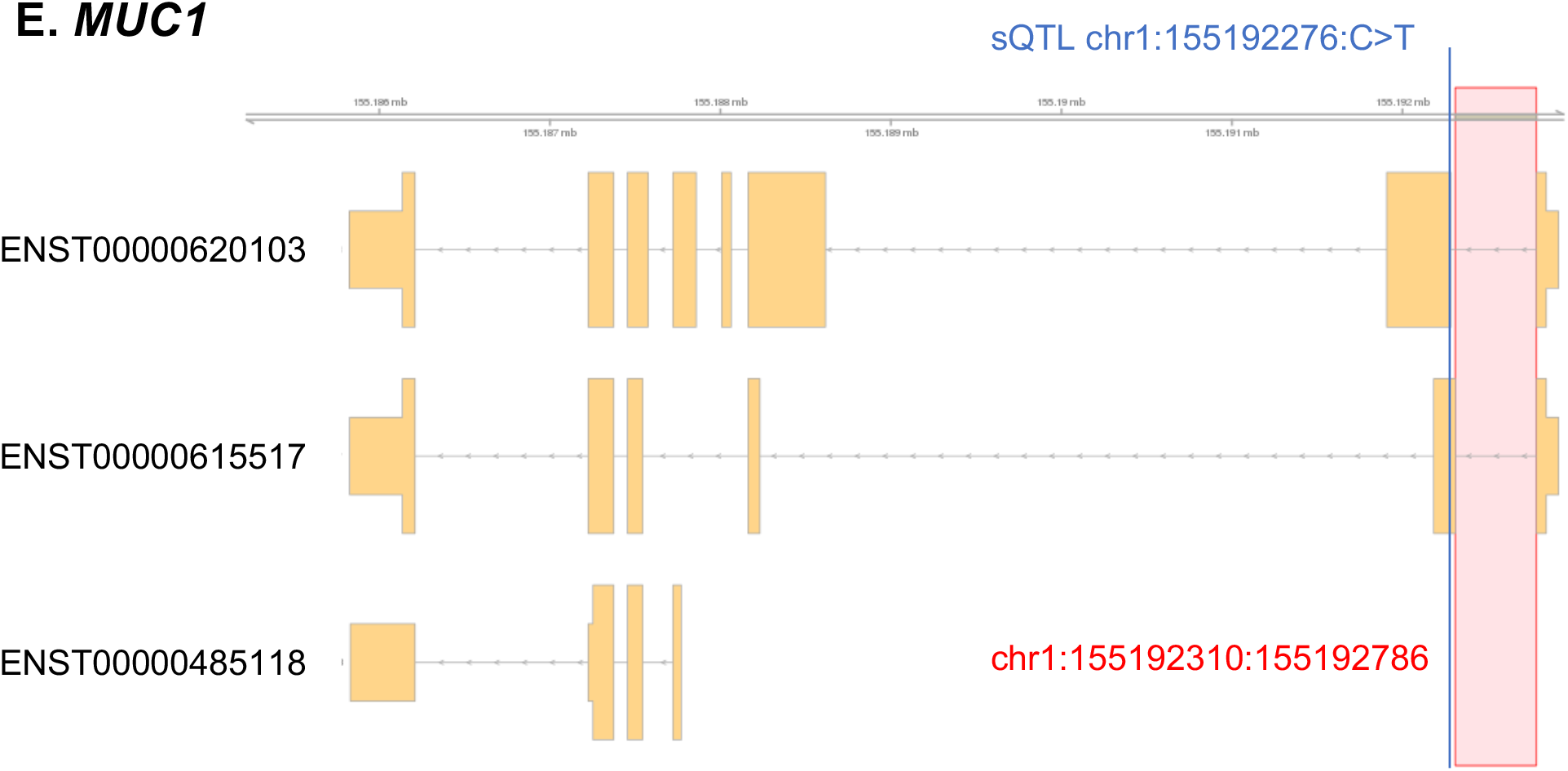
The gene plots. The major gene-coding transcripts were manually curated to visualize, in relation to the sQTLs and the excised introns quantified by LeafCutter. (A) *OAS1* (B) *ATP11A* (C) *DPP9* (D) *NPNT* (E) *MUC1*

## Contributions

Conception and design: TN, and JBR. Formal analysis: TN. Data curation: TN, and YF. Interpretation of data: TN, YF, RJA, BGG, and JBR. Funding acquisition: JBR. Investigation: TN, and YF. Methodology: TN, YF, and JBR. Project administration: JBR. Resources: JBR. Supervision: YF, and JBR. Validation: TN, YF, and JBR. Visualization: TN. Writing—original draft: TN. Writing—review and editing: TN, YF, JW, RJA, BGG, SZ, and JBR. All authors were involved in further drafts of the manuscript and revised it critically for content. All authors gave final approval of the version to be published. The corresponding authors attest that all listed authors meet authorship criteria and that no others meeting the criteria have been omitted. All authors were involved in further drafts of the manuscript and revised it critically for content. All authors gave final approval of the version to be published. The corresponding author attests that all listed authors meet authorship criteria and that no others meeting the criteria have been omitted.

